# Strategies for exiting COVID-19 lockdown for workplace and school: a scoping review protocol

**DOI:** 10.1101/2020.09.04.20187971

**Authors:** Daniela D’Angelo, Daniela Coclite, Antonello Napoletano, Alice Josephine Fauci, Roberto Latina, Laura Iacorossi, Marco Di Nitto, Primiano Iannone

## Abstract

**Objective:** The main objectives of this review are to summarise the existing literature and to identify strategies for exiting lockdown during the COVID-19 pandemic, with a focus on reopening schools and returning to work.

**Introduction:** After months of strict quarantine, several countries are planning exit strategies to progressively lift social restrictions without giving rise to an increase in the number of COVID-19 cases. Although several strategies have been studied in terms of how and when to relax such stringent constraints, there is a lack of consensus on the optimal strategy for managing the pandemic beyond lockdown. The risks posed by delaying return to work and school openings are real and sizeable, particularly for relevant working sectors and for students from low-income families

**Inclusion criteria:** This review will consider studies that focussed on relaxation strategies for lockdown exit among workers and students facing an epidemic /pandemic crisis.

**Methods:** The searches will be conducted across four databases (MEDLINE, EMBASE, SciSearch, Google Scholar), and the bibliography of all selected studies will be hand-searched. In addition, because the topic is new, relevant literature will be checked using daily, updated COVID-19 collections from NCBI (LitCovid) and MedRxiv servers. Studies published in English, German, Spanish, Italian and French will be included, with no limits on publication dates. This review will consider all study designs, regardless of their rigor. The review method will be based on a two-phase approach: a title and abstract screening, and a full-text review performed by two independent researchers. Data will be summarised and categorised, and results will be presented in a tabular/diagrammatic form.

## Background

COVID-19 has infected millions of people worldwide, resulting in a serious health crisis in terms of morbidity and mortality. Additionally, the experience of economically developed countries has demonstrated that COVID-19 can overwhelm healthcare capacity (1–3). Since neither pharmaceutical treatments nor vaccines are available, many countries have implemented strict social distancing measures and introduced lockdowns to prevent the spread of the virus. These measures have been necessary in order to substantially reduce the growth of the epidemic and transmission of the virus, and to reduce the intensity, or peak, of the epidemic (flatten the curve) (4). After months of strict quarantine, social distancing measures and lockdown, governments in various countries, policy decision-makers and citizens are all keen to see a relaxation in restrictions. Relaxation strategies are needed as lockdown has significantly restricted people’s civil rights and their income; indeed, from an economic and social viewpoint confinement measures are not sustainable in the long run, and a well-designed exit strategy is therefore crucial.

One of the main concerns is that such a relaxation might trigger a further epidemic wave: if lockdown is abandoned too early or without restrictions, infection could rebound and successive interventions may become necessary (5). After achieving adequate infection control, strategic public health efforts are directed toward gradually reopening society through constant epidemiological monitoring. The post-pandemic transmission dynamics will also depend on factors such as the intensity and timing of control measures, but also on the degree of seasonal variation and the duration of immunity. Restrictions should be relaxed gradually so that Rt does not exceed 1, otherwise cases would once again increase exponentially, unleashing a second wave of infection (6, 7).

Since person-to person transmission is mostly driven by social interactions, closing schools and asking people to do their jobs from home were among the first decisions taken in many states. Logically, as no vaccine will be available for several months, this pandemic can only be kept under control through major social reorganisation (8), so great attention should be paid to safely managing and reorganising the reopening of schools and the return to work (9–11).

To curb the spread of the virus, physical distancing measures, social hygiene practices and deep cleaning of shared surfaces is highly recommended(12). The work-transition strategy might require gradually sending people at low risk from COVID-19 back to work. Their safety should be guaranteed by using face-coverings and by the testing, tracking and isolation of infected individuals, and screening of asymptomatic individuals (9–11). After the reopening of schools, social distancing will be important in preventing a recurrence of the epidemic, and governments need to establish realistic and practical social distancing policies, and preventive measures against epidemics (6, 13–15). A systematic review suggested that the use of masks in schools can be effective in limiting the transmission of respiratory infections, (16) (17).

Although the COVID-19 pandemic has stimulated clinical and epidemiological studies, as well as mathematical models on how to exit lockdown (6, 13–15), there is currently no scientific consensus on safe conditions for returning to normal life. Planning how and when to lift restrictions is an issue of utmost relevance and the need to get countries back to work and schools is urgent and requires an understanding of the exit strategies taken across the world to date.

There is therefore a need to gather all the knowledge and experience from existing lockdown exit strategies so as to continually improve these programmes and to support new developments. For this purpose, we chose a scoping review, rather than a systematic review, as we expect to find many different study designs that explore the topic at hand. Moreover, our research question will be kept broad to capture the whole range of research on lockdown exit strategies.

Scoping reviews are considered a valid method for reviewing the available literature on a given topic in order to clarify the characteristics, determine the range of studies available, and summarise research results.

The primary objective of this review is to summarise the existing literature on strategies for exiting lockdown during the COVID-19 pandemic, with a focus on reopening schools and returning to work. By identifying the lockdown exit strategies for COVID-19, or for any other similar pandemic, this review attempts to establish a foundational understanding of how these strategies were implemented and to gather the key success factors and recommendations. In the medium term, the review seeks to consider how existing strategies could be used by governments and policy makers.

## Methods

The proposed scoping review follows the framework proposed by Arksey and O’Malley (18), which has been further developed by Levac et al. (19) and the Joanna Briggs Institute (20). According to this method, there are six stages in undertaking a scoping review: (1) identifying the research question; (2) identifying relevant studies; (3) selecting studies; (4) charting the data; (5) collating, summarising and reporting the results and (6) consulting the relevant stakeholders. Although the latter was described by Arksey and O’Malley as optional but desirable, we will not be able to conduct a stakeholder consultation due to time constraints.

### Stage 1: Identifying research questions

The objective of this scoping review is to map the accessible research literature to answer the research question: Which non-pharmacological lockdown exit strategies for workers and students during the COVID-19 or any other similar pandemic have been reported in literature?

Although this question is broad enough to ensure that all the existing literature is captured and analysed in the review, consultation with the research team could be resulted in identifying other sub questions to guide the subsequent stages of the scoping review.

### Stage 2: Identifying relevant studies

The search strategy was iteratively developed by the research team in collaboration with an experienced medical librarian. A preliminary search, run on May 20th, 2020 was performed on MEDLINE to identify index terms and keywords. This search strategy was translated and tailored for use in biomedical databases (EMBASE, SciSearch) and in online grey literature databases (Google Scholar) through CASSTN system. The mentioned databases were searched from inception until May 25th, 2020.

An initial scan of biomedical databases demonstrated that due to the newness of the topic, the databases selected were not likely to identify results related to the focus of this scoping review. To overcome this and ensure that the scoping review is comprehensive and up-to-date, the research team will also extensively hand-search various reference lists of included studies and key journals. National and international government policy documents will also be monitored. Manual searches of the literature will be undertaken using daily updated COVID-19 collections from the National Centre for Biotechnology Information (https://www.ncbi.nlm.nih.gov/research/coronavirus/docsum?filters=topics.Prevention), MedRxiv (https://www.medrxiv.org/), and BioRxiv (https://www.biorxiv.org/) preprint servers. Specifically, the latter two sources will be searched until July 1, 2020.

The complete and definitive search strategy for MEDLINE can be found in the online supplementary Appendix I; further search strategy details across bibliographic databases are available upon request from the first author.

### Stage 3: Study selection

This scoping review will adopt the following inclusion criteria: (1) studies must refer to workers or students of all ages facing an epidemic/pandemic crisis; (2) articles that measure or discuss relaxation strategies to exit lockdown (mass quarantine) during an epidemic/pandemic crisis, (3) quantitative studies of any study design can be included (i.e. randomised controlled trials, cohort, case–control, quasi-experimental, cross-sectional, mathematical model), as well as editorials, letters, and commentaries.

Studies will be excluded from the scoping review if they deal with an epidemic/pandemic crisis in the presence of a vaccine or herd immunity, and if they considered the health care workers as study population.

Searches will be limited to articles in English, Italian, German, French and Spanish, with no limits on publication dates.

While these inclusion and exclusion criteria will remain strict, we will also adopt an iterative approach to define additional, specific criteria as the research team becomes more confident with the topic through a full review of potential studies.

Search results will be imported to EndNote X9 (Clarivate Analytics, PA, USA) for screening and further reviewing. During this process, duplicates will be identified and removed. Studies will be independently screened by two reviewers to identify inclusion and exclusion criteria. Grey literature will be title-screened for all the resulting pages in the search. The review method will be based on a two-phase approach: a title and abstract review, and a full-text review, performed by two independent researchers. Any discrepancies will be discussed between the two reviewers until consensus is reached. In the event of any disagreement, this will be discussed in detail with a third reviewer. Reasons for excluding studies after a full-text review will be documented. Moreover, full-text studies excluded from the final version of this scoping review will be provided in an appendix. Data extraction from included studies will then be provided.

The reporting method of this review follows the Preferred Reporting Item for Systematic Reviews and Meta-Analyses Scoping Review (ScR) Checklist recently developed for scoping reviews (21). The study selection process will be summarised using a PRISMA flow diagram (16).

### Stage 4: Data charting process

The standard bibliographical information (authors, journal and year of publication, where the study was published/conducted) will be provided. For each article, information on the objective and the interventions covered—concept, context, characteristics of the study populations, length and intensity of the interventions, types of outcomes assessed, key findings—will be presented in a table.

To answer the research question, a first data extraction tool draft adapted from the standardised JBI data extraction tool will be used (table 1). The data extraction tool will be further refined when the research team has gained greater awareness of the topics in the included studies. The data extraction form will be pilot tested by two team members on a sample of five articles in order to ensure that the coding has been reliably applied. Any modifications will be detailed in the full scoping review.

**Table 1.**
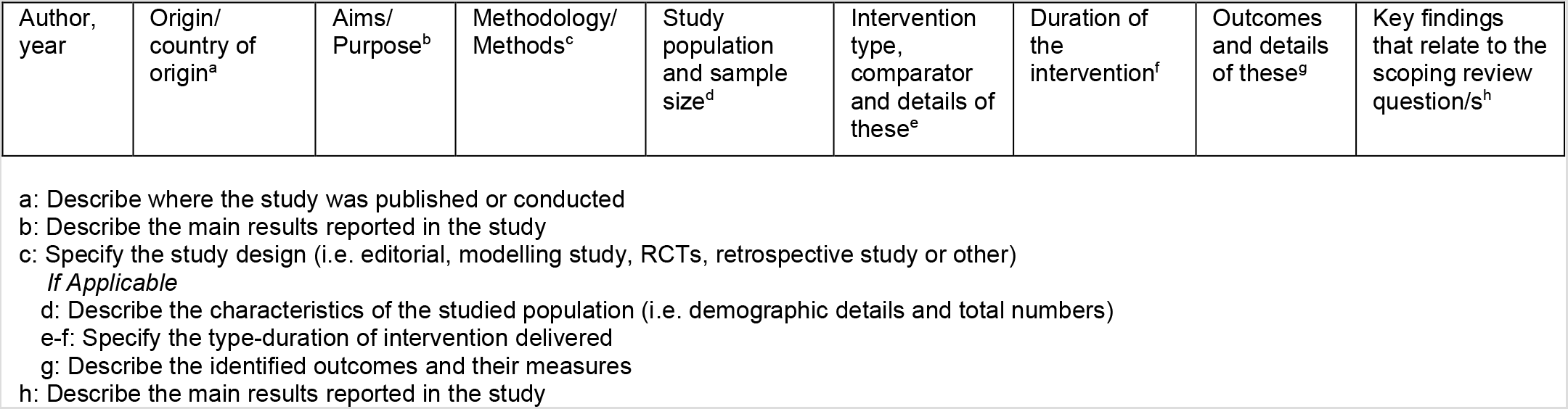
Preliminary data extraction tool and its description

#### 4.1 Assessment of Methodological Quality

In accordance with Arksey and O’Malley (18) no assessment of the methodological quality of the included papers will be conducted. Details of article type and methodology will be reported in a summary table to provide context of the maturity of the evidence, and the strengths, weaknesses and applications of different methodologies will be discussed. However, due to the newness of the topic we are likely to include many preliminary mathematical models not yet peer reviewed, therefore, to avoid drafting inconclusive and biased results a quality assessment of such studies will be performed.

### Stage 5: Summarizing results

The extracted data will be presented in diagrammatic or tabular form in a manner that aligns with the objective of this scoping review. Given that we expect varying designs and populations across the different studies that will be eligible for this scoping review, the analysis of the results will depend on the type of data that is collected. Data will be examined analytically (i.e. summarising the frequency/demographics of studies and study populations) and narratively (i.e. giving a narrative summary of the individual study results for each identified topic). In order to present the data in a comprehensive and useful manner, within the two main categories of workers and students data will be sub-divided into new emerging categories such as; school level (i.e. primary, secondary), workplace (i.e. office, factory), and type of occupation (i.e. business, hospitality, education).

The topics of the retrieved articles will probably be classified according to the type of studies from which they derived (clinical-epidemiological studies, and mathematical models).

## Data Availability

This is a scoping review protocol so no data are included in the manuscript

## Appendix I. Search strategy

Search conducted on MEDLINE on 25 May 2020

| Search | Query | Record retrieved |
| --- | --- | --- |
| #1 | COVID 19/CT,TI,AB OR COVID19/CT,TI,AB | 14,462 |
| #2 | SARS COV2/CT,TI,AB | 175 |
| #3 | SARS COV 2/CT,TI,AB | 3,794 |
| #4 | NCOV/BI OR N COV/CT,TI,AB | 747 |
| #5 | NOVEL CORONAVIRUS/CT,TI,AB OR NOVEL CORONA VIRUS/CT,TI,AB | 2,327 |
| #6 | NEW CORONAVIRUS/CT,TI,AB OR NEW CORONA VIRUS/CT,TI,AB | 342 |
| #7 | CORONAVIRUS DISEASE#/CT,TI,AB OR CORONA VIRUS DISEASE#/CT,TI | 3,513 |
| #8 | SEVERE ACUTE RESPIRATORY SYNDROME CORONAVIRUS 2/CT,TI,AB | 4,283 |
| #9 | SEVERE ACUTE RESPIRATORY SYNDROME CORONA VIRUS 2/CT,TI,AB | 10 |
| #10 | PANDEMIC/CT,TI,AB | 33,361 |
| #11 | PANDEMIC/CT,TI,AB | 5,300 |
| #12 | PANDEMIA#/CT,TI,AB | 377 |
| #13 | INFLUENZA, HUMAN/CT,TI,AB | 48,754 |
| #14 | INFLUENZA/BI AND HUMAN#/BI | 81,132 |
| #15 | #1 OR #2 OR #3 OR #4 OR #5 OR #6 OR #7 OR #8 OR #9 OR #10 OR #11 OR #12 OR #13 OR #14 | 108,878 |
| #16 | FACEMASK#/CT,TI,AB | 1,242 |
| #17 | FACE# MASK#/TI,AB | 3,123 |
| #18 | MASS GATHER?/CT,TI,AB | 645 |
| #19 | RETURN TO WORK/CT,TI,AB | 9,832 |
| #20 | RETURN?(S)WORK?/TI,AB | 15,549 |
| #21 | BACK TO WORK/TI,AB | 568 |
| #22 | SEROEPIDEMIOLOGIC STUDIES/CT,TI,AB | 20,462 |
| #23 | SEROEPIDEMIOLOGIC? STUD?/TI,AB | 1,578 |
| #24 | MASS SCREENING+NT/CT | 127,032 |
| #25 | MASS SCREENING/TI,AB | 4,989 |
| #26 | SOCIAL DISTANCE/CT,TI,AB | 3,449 |
| #27 | SOCIAL DISTAN?/TI,AB | 1,643 |
| #28 | LOCKDOWN? | 402 |
| #29 | LOCK DOWN? | 51 |
| #30 | CONTACT TRACING/CT,TI,AB | 5,296 |
| #31 | CONTACT TRACKING/CT,TI,AB | 11 |
| #32 | REOPEN?/CT,TI,AB OR RE OPEN?/CT,TI,AB | 4,279 |
| #33 | (SCHOOLS+NT/CT OR SCHOOL#/TI,AB) | 327,111 |
| #34 | #32 AND #33 | 76 |
| #35 | #15 AND (#16 OR #17 OR #18 OR #19 OR #20 OR #21 OR #22 OR #23 | 2,410 |
|  | OR #24 OR #25 OR #26 OR #27 OR #28 OR #29 OR #30 OR #31 OR #34) |  |
| #36 | #35 AND (ENGLISH/LA OR ITALIAN/LA OR FRENCH/LA) | 2,304 |
| #37 | #35 AND (SPANISH/LA OR GERMAN/LA) | 36 |
| #38 | #36 OR #37 | 2,336 |

